# A novel reperfusion method in patients with acute ST elevation myocardial infarction during primary percutaneous coronary intervention: A randomized control trial

**DOI:** 10.1101/2023.10.13.23297032

**Authors:** Ji-Fang He, Yi-Xing Yang, Yu Liu, Zong-Sheng Guo, Hao Sun, Lu Liang, Tao Jiang, Qi Yang, Xiang-Min Lin, Mu-Lei Chen, Pi-Xiong Su, Xin-Chun Yang, Li Xu, Le-Feng Wang

**Affiliations:** Heart Center and Beijing Key Laboratory of Hypertension, Beijing Chaoyang Hospital, Capital Medical University, No. 8, Gongti South Road, Chaoyang District, Beijing, 100020, China; Department of Radiology, Beijing Chaoyang Hospital, Capital Medical University, No. 8, Gongti South Road, Chaoyang District, Beijing, 100020, China

**Author notes:** These authors contributed equally to this study (Ji-Fang He, Yi-Xing Yang). Corresponding Author: Li Xu, MD; Le-Feng Wang, professor. Heart Center and Beijing Key Laboratory of Hypertension, Beijing Chaoyang Hospital, Capital Medical University, No. 8, Gongti South Road, Chaoyang District, Beijing, 100020, China.

## Abstract

**BACKGROUND:** Reperfusion injury is associated with larger infarct size and drawback of long-term outcome in patients with acute ST-segment elevation myocardial infarction (STEMI) undergoing primary percutaneous coronary intervention (PPCI). Volume-Controlled Reperfusion (VCR) is our original reperfusion method combined with accurate gentle reperfusion and active protection for unstable plaque. A single center, prospective, randomized clinical trial was conducted to demonstrate the feasibility, clinical safety, and potential effects of VCR in STEMI patients during PPCI.

**METHODS AND RESULTS:** A total of 60 patients with STEMI presenting within 12 h of symptom onset undergoing PPCI were randomized enrolled to VCR group (n=30) or control group (n=30). Patients in VCR group recanalized with VCR followed by routine PCI. Patients in control group recanalized with routine PCI procedure only. The primary endpoint was all-cause death and major adverse cardiac events (MACE) during hospitalization and followed up, the main second endpoints were MVO extent measured 5-7 days post PPCI and left ventricular function. Baseline characteristics were well matched between two groups. No MACE from in-hospital to 1 year followed up in VCR group. In-hospital echocardiography based left ventricular eject fraction (LVEF) in VCR group compared with control group was (57.6 ± 6.2) % vs (52.9 ± 8.5) %, p = 0.018; especially in anterior STEMI VCR subgroup (55.1 ± 6.3) % v (s 48.6 ± 6.4) %, p = 0.003. There was no significant difference in infarct size, MVO mass or MVO mass/infarct mass ratio by late gadolinium enhancement-cardiac magnetic resonance imaging (LGE-CMR). In hospital CMR based LVEF in anterior STEMI VCR subgroup was (50.55 ± 7.55) % vs (40.53 ± 7.39) %, p = 0.002. Follow up echocardiography based LVEF were (64.12 ± 6.35) % vs (59.44 ± 8.20) %, p = 0.025 between two groups and (62.07 ± 7.06) % vs (55.94 ± 6.70) %, p = 0.016 between anterior STEMI subgroups.

**CONCLUSIONS:** This is the pilot RCT study demonstrated that VCR method was feasibility and clinical safe among patients with acute STEMI during PPCI to 12-month follow-up, indicated potential attenuate effect in reperfusion injury as well. Staged II RCT is needed to verify clinical benefit.

**REGISTRATION:** URL: http://www.chictr.org.cn. Unique identifier: ChiCTR00052856

## CLINICAL PERSPECTIVE

**What Is New?**

- This method, Volume-Controlled Reperfusion (VCR), for the first time provides accurate quantitative gentle reperfusion in primary PCI in Cath-lab setting. VCR integrates advantage of different procedures to attenuate reperfusion injury, such as gentle reperfusion, active plaque protection and intermittent blood flow control.
- Our findings demonstrate that VCR is clinically safe and feasible with improved left ventricular function.

**What Are the Clinical Implications?**

- VCR might be a promising procedure to attenuate reperfusion injury in patients with ST-segment elevation myocardial infarction undergoing primary percutaneous coronary intervention.
- VCR might be given to different clinical scenario, such as pulmonary or cerebral embolism.
- VCR might provide a reference for pharmacological conditioning medication delivery in the future.

## Introduction

Ischemic reperfusion injury (IRI) prevention is a challenging task during primary percutaneous catheterization intervention (PPCI) in STEMI patients. Microvascular obstruction is an important pathophysiological index and procedure of IRI and myocardium damage and repair, considered to be predictor of in-hospital MACE and long-term outcome in patients post-PPCI with acute STEMI. Recently MVO become a surrogate endpoint in clinical trial. ^1, 2^ Many treatments, aimed to rescue cardiomyocyte in aera at risk (AAR) and myocardium salvage, have been transferred from experiment to bedside but failed show significant clinical benefit in stage III randomized controlled trial (RCT), such as ischemia post conditioning (IPost), controlled hypothermia reperfusion etc. ^3^

Controlled gentle reperfusion aimed to avoid abrupt blood flow has been investigated extensively in coronary ischemia and has consistently been shown to attenuate reperfusion injury. However, most studies of gentle reperfusion were limited in cardiopulmonary bypass or animal studies because of difficulty to control blood flow in clinical setting during PPCI. ^4^

We proposed “volume-controlled reperfusion (VCR) method” for quantitative control of blood flow during PCI procedure. This method combined with advantage of accurate gentle reperfusion, IPost-like intermittent blood flow, and active protection for unstable plaque. (Figure 1) VCR is composed of two key procedures, Firstly, set up “intra-coronary faucet” to control blood flow. Prophylactically suspend antegrade flow with seal balloon on culprit lesion, followed with an aspiration catheter positioned distal from balloon tip based on dual-catheter technique. Secondly, manual blood reperfusion via aspiration catheter with predetermined perfusion volume. Routine stent or drug coated balloon (DCB) as usual after VCR.

**Figure 1.**
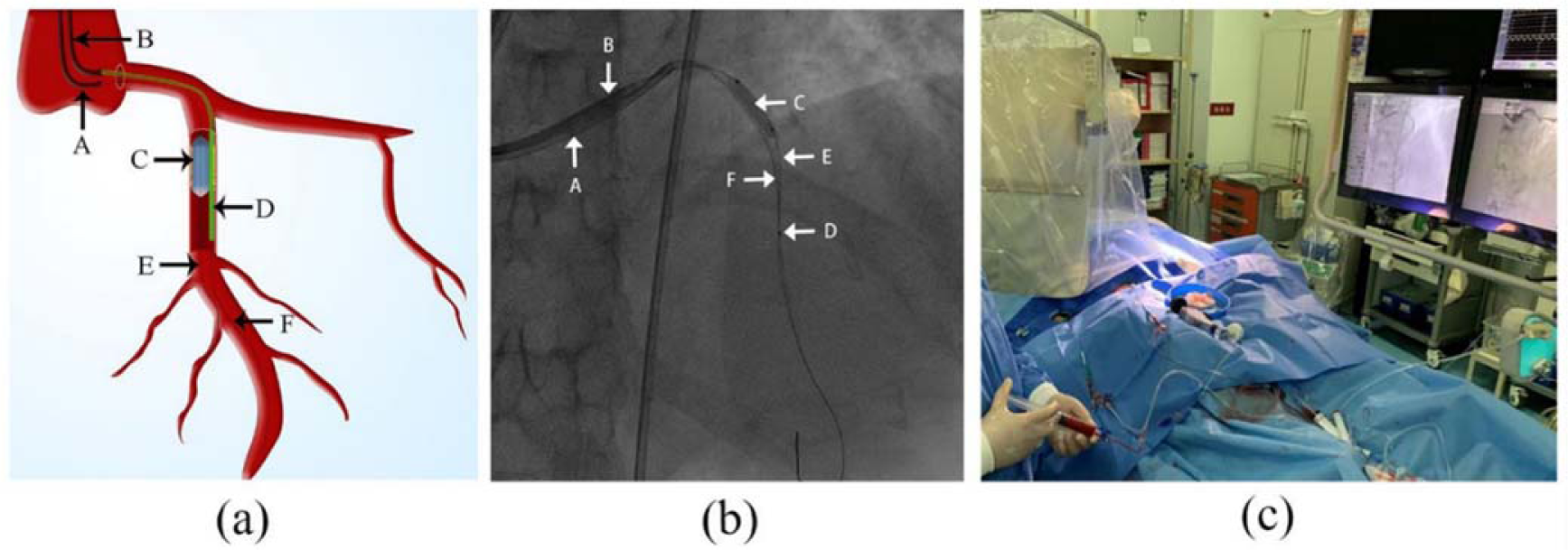
Illastration of Volume-Controlled Reperfusion Method. (a) VCR schematic Illustration: (b) Screen shot of VCR Illustration: (c) VCR Method in Cath-lab Practice: A. The radial artery route guide catheter 1: B. The femoral artery route guide catheter 2; C. Sealing balloon suspend forward blood flow: D. Aspiration catheter: E. Work horse guide wire 1: F. Work horse guide wire 2

We therefore performed a proof-of-concept single center RCT to evaluate feasibility and safety issue of VCR in patients with STEMI (STEMI VCR), potential effect of IRI attenuation was under observation.

## Methods Patients

Patients with symptoms consistent with acute STEMI less than 12 h, with ≥2mm of ST segment elevation in ≥2 contiguous leads, angiography indicated proximal de novo occlusion lesion in the target vessel were eligible for enrollment. The inclusion criteria were age between 18 and 80 years; first STEMI attack; symptom to onset <12 hours; occlusion at the proximal or mid region on angiography; Thrombolysis in Myocardial Infarction (TIMI) grade 0 or I flow in the culprit vessel. The exclusion criteria were previous myocardial infarction; previous coronary bypass surgery; clinically unstable (hemodynamically unstable, shocked, unconscious patients) mechanical complications; severe diffused multivessel disease; distal culprit lesion or in non-dominant vessel.

### Study Design and Protocol

The study was conducted in accordance with the Declaration of Helsinki and the protocol was approved by the ethics committee in Beijing Chao-Yang Hospital, Capital Medical University.

The loading dose of aspirin 300 mg and clopidogrel 300 mg given before procedure. Once diagnostic angiography showing proximal occlusion of a dominant coronary artery and fulfill the inclusion criteria, patients were asked for oral informed consent by catheterization table. Informed written consent was obtained within 24 hours. Intravenous bolus of heparin was 70-100U/kg. Intravenous glycoprotein IIb/IIIa inhibitor or intra-aortic balloon pumping (IABP) was left to operator discretion.

Patients were randomized into VCR group or control group according to computer-generated random sequence. Patients in VCR group started with VCR followed by routine PCI. Patients in control group recanalized with routine PCI procedure only. No specific restriction for manual thrombus aspiration/AngioJet^®^(Bayer HealthCare) thrombectomy system and/or pre-dilate before stenting in routine PCI group. TIMI III blood flow were observed during PPCI procedure. Residual thrombus was estimated based on Thrombolysis In Myocardial Infarction (TIMI) thrombus grade (TTG) scoring system after PTCA and before stent deployed.^5^

A 12-lead ECG was obtained pre- and 60min post-procedure. Bedside echocardiography, hemodynamic and ECG monitoring in cardiac care unit post PPCI. Infarct size (IS) and MVO were assessed by late gadolinium enhancement-cardiac magnetic resonance imaging (LGE-CMR) at day 5-7 post PPCI. Echocardiography and LGE-CMR study were blinded to the randomized treatment.

All patients were treated with dual anti platelet therapy for at least 9-12 month. Statins, beta-blockers, and angiotensin-converting enzyme inhibitors or receptor blockers were prescribed in the absence of contraindications. Clinical follow-up was performed from 30 days to 12 months by routine outpatient clinic visits or telephone interviews. Echocardiography was carried out on the last follow up.

### VCR Method Description

VCR method was evaluated in our initial feasibility study.^6^ Basically, (Figure 1) catheter engaged to coronary orifice with dual-catheter technique via radial and femoral artery approach respectively, workhorse wire A and B were manipulated to distal vessel from each guide catheter respectively; forward blood flow was sealed with 3.0/3.5mm NC balloon (according to reference vessel diameter close to 1:1 ratio) on culprit lesion at 6-8 atm; aspiration catheter was advanced 10-15mm distal from NC balloon tip followed with gentle contrast puff via aspiration catheter to prove distal part patency; finally, intra-aspiration catheter infusion of mixture solution (artery blood 10ml+ heparin NS 10ml) at 20ml/min for 10 rounds, adjust volume and speed according to blood pressure and heart rate variation. Then, deflate NC balloon, manual aspiration during aspiration catheter withdraws. Followed by stent or DCB as routine PCI procedure.

### CMR Analysis

Cardiac MRI was performed at 3T scanner (Prisma, Siemens Healthcare, Erlangen, Germany) for CMR imaging of cardiac function and tissue contrast enhancement by using an anterior phased-array body coil (18-element) and a posterior phased-array spine coil (24-element). After acquisition of scout images, retrospective electrocardiographic gating cine imaging was performed using a segmented balanced steady-state free precession sequence in continuous short-axis views, spanning the entire LV from base to apex. After the acquisition of cine images, gadolinium-based contrast medium was administered intravenously at 0.1 mmol/kg body weight. LGE images were obtained in short-axis locations from the base to the apex of the LV, by using a three-dimensional inversion recovery T1 turbo field-echo sequence, 10 minutes after contrast administration. The MR images were analyzed at CVI software (cvi42, Circle Cardiovascular Imaging Inc., Calgary, Canada) by 2 radiologists blinded to patient data. To assess IS, in all short-axis images the myocardial contours and late-enhancement boundaries were traced manually. MVO was defined as the presence of hypo-intense areas within the infarct core scar characterized by hyper enhanced myocardium.

### Endpoints

The primary endpoint was all-cause death during hospitalization, major adverse cardiac event (MACE) during in-hospital and the long-term follow up. The second endpoints included TIMI grade after PCI; infarct size and microvascular obstruction (MVO) assessed by LGE-CMR; left ventricular ejection fraction (LVEF) measured by echocardiography and CMR; Cardiac biomarkers peak level after PCI.

MACE was defined as the composite of cardiac death, myocardial reinfarction, target vessel revascularization (TVR) and heart failure. Definitions of cardiac death, myocardial reinfarction and TVR corresponded with the Academic Research Consortium criteria. Heart failure was defined as any congestive heart failure.

### Statistical Analysis

Intent-to-treat (ITT) principle was adopted for data analysis in this experiment. In addition, the “Per-Protocol (PP) analysis” and “AT-as-treated analysis” were performed. SPSS 26 software was used or data analysis, and the measurement data conforming to normal distribution were marked with mean ±standard deviation (SD) indicates that the T test is applied to the comparison between groups to the normal distribution. The measurements were compared between the median and interquartile ranges (IQR) to those does not conform normal distribution with the Mann-Whitney test. Counting data is expressed in percentiles, and comparison between groups is performed using chi-square tests or Fisher’s exact tests. The cumulative event rate of MACE will be estimated by Kaplan-Meier method and log-rank tests were performed for comparison between groups. Variables with P < 0.05 were considered statistically significant.

## Results

257 patients with STEMI were screened for study eligibility in Beijing Chao-Yang Hospital between November 2021 to July 2022. 60 patients were randomized to VCR group (n =30) and control group (n =30). A total of 17 patients did not complete the CMR examination, 9 in VCR group and 8 in control group (Figure 2). A total of 43 patients in both groups completed LGE-CMR.

**Figure 2.**
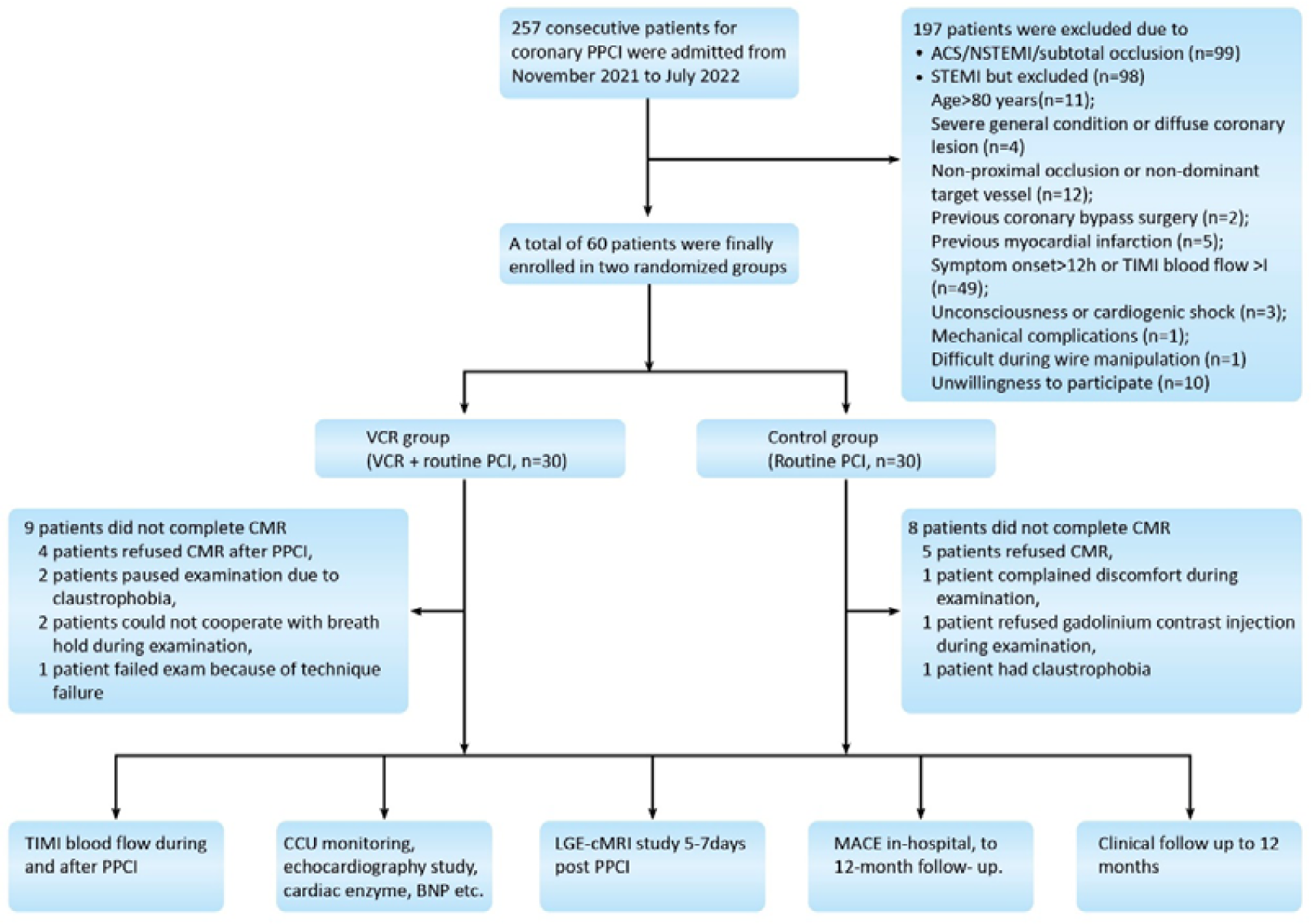
Flow Chart of STEMI VCR Study. A total of 60 patients were enrolled, all patients were completed 12-month follow up, 17 patients did not complete CMR study. STEMI indicates ST elevated myocardial infarction; ACS, acute coronary syndrome; NSTEMI, non-ST elevated myocardial infarction; VCR, volume-controlled reperfusion; CMR, cardiac magnetic resonance; PPCI, primary percutaneous coronary intervention; TIMI, Thrombolysis in Myocardial Infarction; CCU, cardiac care unit; BNP, B-type natriuretic peptide; LGE-cMRI, late gadolinium enhancement-cardiac magnetic resonance imaging; MACE, major adverse cardiac events.

### Baseline Characteristics

Baseline clinical and angiographic features were well matched between the 2 groups (Tables 1-2). The culprit vessel included the left anterior descending coronary artery in 37 cases (61.7%) of patients, 20 cases (51.3%) in right coronary artery, and the left circumflex coronary artery were in 3 cases (8.3%). Single lesion in main branch was treated in all patients. Aspiration was performed in 1 patient in VCR group (3.3%) and 7 cases (11.7%) in control group. Balloon pre-dilation before stent implantation was performed in control group with a median 2.0-mm diameter balloon. Use of antiplatelet agents, anticoagulants, and other study procedures were also similar between the 2 groups. Non target vascular revascularization (0.0%) in VCR group, 1 patient (3.3%) in control group underwent target vascular revascularization in LAD.

**Table 1.** Baseline Characteristics and Blood Test Results of the Study Patients.

|  | VCR Group<br>(n=30) | Control Group (n=30) | P value |
| --- | --- | --- | --- |
| Age, yrs | 56.83 ± 12.48 | 59.90 ± 10.24 | 0.30 |
| Male | 21 (70.0) | 25 (83.3) | 0.22 |
| Body mass index, kg/m<sup>2</sup> | 26.20 ± 3.47 | 24.86 ± 2.78 | 0.10 |
| Hypertension | 18 (60.0) | 19 (63.3) | 0.79 |
| Diabetes mellitus | 9 (30.0) | 10 (33.3) | 0.78 |
| Hyperlipidemia | 5 (16.7) | 7 (23.3) | 0.52 |
| Chronic obstructive lung disease | 0 (0.0) | 1 (3.3) | 1.00 |
| Previous Stroke | 1 (3.3) | 2 (6.7) | 1.00 |
| Cigarettes smoking | 22 (73.3) | 20 (66.7) | 0.57 |
| Family history of cardiovascular disease | 22 (73.3) | 20 (66.7) | 0.57 |
| On admission systolic pressure, mmHg | 130.1 ± 19.7 | 128.3 ± 19.7 | 0.52 |
| On admission diastolic pressure, mmHg | 80.7 ± 14.4 | 78.5 ± 14.6 | 0.29 |
| On admission heart rate, bpm | 82.1 ± 12.4 | 80.4 ± 17.8 | 0.46 |
| LVEF, % | 58.8 ± 10.3 | 60.0 ± 10.4 | 0.39 |
| TNI, ng/ml | 2.64 (0.11, 134.07) | 0.41 (0.10, 25.38) | 0.35 |
| CKMB, ng/ml | 19.03 (2.03, 99.85) | 4.95 (2.68, 50.30) | 0.48 |
| BNP, pg/ml | 46.00 (23.25, 177.50) | 49.00 (27.75, 184.75) | 0.63 |
| ALB, g/l | 43.04 ± 4.25 | 43.64 ± 2.94 | 0.52 |
| GLB, g/l | 28.05 ± 5.27 | 28.55 ± 3.99 | 0.68 |
| TC, mmol/l | 4.69 ± 0.95 | 5.17 ± 0.94 | 0.05 |
| TG, mmol/l | 2.53 ± 2.14 | 1.95 ± 1.30 | 0.21 |
| HDL-C, mmol/l | 0.95 ± 0.25 | 1.03 ± 0.17 | 0.15 |
| LDL-C, mmol/l | 3.13 ± 0.87 | 3.77 ± 0.91 | 0.01 |
| LPA, mg/dl | 14.95 (7.00,26.00) | 19.75 (13.30,33.98) | 0.09 |
| GLU, mmol/l | 9.61 ± 4.01 | 9.52 ± 3.31 | 0.93 |
| CR, umol/l | 75.27 ± 19.14 | 71.53 ± 14.65 | 0.40 |
| WBC, ×10<sup>9</sup>/l | 10.55 ± 3.47 | 10.98 ± 3.61 | 0.63 |
| HGB, g/l | 148.63 ± 18.20 | 149.97 ± 16.64 | 0.77 |
| PLT, ×10<sup>9</sup>/l | 239.30 ± 91.29 | 227.13 ± 65.57 | 0.56 |
| D-dimer, mg/l | 0.24 (0.14,0.42) | 0.25 (0.19, 0.50) | 0.43 |
| FIB, mg/dl | 300.66 ± 70.26 | 290.34 ± 68.29 | 0.57 |

VCR: volume-controlled reperfusion; TNI: troponin I; CK-MB: creatine kinase isoenzyme MB ; BNP: brain-type natriuretic peptide; ALB: albumin; GLB: globulin; TC: total cholesterol, ; TG: triglyceride; HDL: high-density lipoprotein cholesterol; LDL: low-density lipoprotein cholesterol; LPA: lipoprotein; GLU: glucose; CR: Creatinine; WBC: white blood cells; HGB: hemoglobin; PLT: platelet; FIB: fibrinogen

**Table 2.** Procedural Details and Medications.

|  | VCR Group<br>(n=30) | Control Group<br>(n=30) | P Value |
| --- | --- | --- | --- |
| Time interval, minutes |  |  |  |
| Chest pain to radial artery sheath | 306.0 ± 134.0 | 298.0 ± 128.5 | 0.81 |
| Total PCI procedure | 43.8 ± 5.4 | 28.2 ± 6.8 | 0.00 |
| Culprit vessel |  |  |  |
| LAD | 17 (56.7) | 20 (66.7) | 0.43 |
| RCA | 12 (40.0) | 8 (26.7) | 0.27 |
| LCX | 1 (3.3) | 2 (6.7) | 1.00 |
| TIMI blood flow pre PCI |  |  | 0.61 |
| 0 | 29 (96.7) | 27 (90.0) |  |
| I | 1 (3.3) | 3 (10.0) |  |
| Stent implantation (DES) | 12 (40.0) | 25 (83.3) | 0.00 |
| DES total (n=37) | 2.0 (2.0, 2.75) | 1.0 (1.0, 1.5) | 0.00 |
| DES total length, mm (n=37) | 48.5 (37.8, 59.8) | 26.0 (22.0, 36.0) | 0.01 |
| DES minimum diameter, mm (n=37) | 3.0 (3.0, 3.5) | 3.0 (3.0, 3.5) | 0.40 |
| Thrombus aspiration | 1 (3.3) | 6 (20.0) | 0.10 |
| IABP | 8 (26.7) | 3 (10.0) | 0.18 |
| GP IIb/IIIa receptor antagonist | 0 (0.0) | 3 (10.0) | 0.24 |
| Combined Non-IRA stenosis > 70% | 15 (50.0) | 17 (56.7) | 0.61 |
| Complete revascularization during the same period (n=32) | 1 (6.7) | 4 (23.5) | 0.34 |
| MBG III | 24 (80.0) | 23 (76.7) | 0.754 |
| CK-MB Peak, ng/mL | 115.0 (61.8, 194.5) | 122.5 (47.2, 170.0) | 0.865 |
| TNI Peak, ng/mL | 134.5 (70.3, 181.0) | 130.1 (61.7, 336.8) | 0.982 |
| 1hr ST-segment resolution | 23 (76.7) | 21 (70.0) | 0.559 |
| Day 3 Echocardiography LVEF, % | 57.6 ± 6.2 | 52.9 ± 8.5 | 0.018 |
| Discharge medications |  |  |  |
| Aspirin | 29 (96.7) | 29 (96.7) | 1.00 |
| ADP antagonists Clopidogrel | 27 (90.0) | 25 (83.3) | 0.71 |
| ADP antagonists Ticagrelor | 3 (10.0) | 5 (16.7) | 0.71 |
| Statins | 28 (93.3) | 26 (86.7) | 0.39 |
| Beta-blockers | 17 (56.7) | 19 (63.3) | 0.79 |
| ACE Inhibitors/ARB | 20 (66.7) | 20 (66.7) | 1.00 |

VCR: volume-controlled reperfusion; PCI: percutaneous coronary intervention; LAD: left anterior descending coronary artery; RCA: right coronary artery; LCX: left circumflex coronary artery; TIMI: thrombolysis in myocardial infarction; DES: drug-eluting stent; IABP: intra-aortic balloon pump; GP: glycoprotein; Non-IRA: non-infarct-related artery; ACEI/ARB: angiotensin converting enzyme inhibitors/angiotensinogen type II receptor blockers.

### VCR method performance

No VCR technique failure except for 1 patient transferred to routine PCI group (difficult in wire manipulation, supported with micro-catheter and multiple balloon pre-dilations). No complications in femoral artery puncture site closured with Perclose ProGlide^®^(Abbott Vascular). 1 patient admitted for emergency thrombolytic therapy in right tibial artery embolism 18 month after PPCI. No air embolus, severe dissection or chunk of thrombus dislocation visualized in culprit artery during VCR procedure.

The average time course from symptom onset to radial artery puncture showed no significant difference between 2 groups (p = 0.81, Table 2). The PCI duration from radial artery puncture to the whole procedure completion was 36.0 ± 9.9 minutes in all patients, of which 43.8 ± 5.4 minutes in VCR group and 28.2 ± 6.8 minutes in control group (p < 0.01). The average VCR set up duration was 10.50±2.80 minutes including femoral artery puncture, sheath insertion, guiding catheter engagement, aspiration catheter position and distal angiography via aspiration catheter.

### Endpoint result Clinical outcomes

There were no deaths or recurrent myocardial infarction in both VCR and control group in-hospital, or 12 month follow up. In control group, there was 4 patients experienced blood pressure drop once recanalization in culprit lesion, two cases in LAD (one of them experienced hypotensive shock need IABP implantation), one case in LCX and two cases in RCA respectively. 1 patient had pericardial effusion 3 hours after PPCI in LAD (emergency pericardial drainage and angiography found no coronary artery rupture); 1 patient with proximal occlusion in LAD experienced ventricular fibrillation 24 hours post PPCI, electrical defibrillation was successful. 2 patients documented severe arrythmia n VCR group during in-hospital monitoring, 1 patient had short-paroxysmal ventricular tachycardia; 1 case of inferior STEMI had a pacemaker implanted for sustained III° AVB; 1 patient’s creatinine was 155μmol/L on admission and was 122 μmol/L in followed up.

### Angiography and STR assessment

No significant difference on post PPCI peak CK-MB or TnI. The 1-hour ST segment resolution rates (STR) were 23 (76.7%) and 21 (70.0%), p = 0.559. TIMI III blood flow were observed in most of cases, except 3 cases in control group. Final TIMI III blood flow cases were 30 (100.0%) and 27 (90.0%), p = 0.237; and final MBG III cases 24 (80.0%) and 23 (76.7%), p = 0.754, in VCR group and control group respectively.

### Echocardiography in-hospital and 12-month follow up

Echocardiography based LVEF on day 3 post PPCI were (57.6 ± 6.2) % and (52.9 ± 8.5) %, p = 0.018, between VCR group and control group. In anterior wall myocardial infarction subgroup study, day-3 LVEF were (55.1 ± 6.3) % and (48.6 ± 6.4%) %, p = 0.003; while no significant difference in LVEF between inferior wall myocardial infarction subgroups (61.25 ± 3.93) % and (61.60 ± 4.57%) %, p = 0.849. Follow up echocardiography based LVEF were (64.12 ± 6.35) % v (s 59.44 ± 8.20) %, p = 0.025 between two groups and (62.07 ± 7.06) % v (s 55.94 ± 6.70) %, p = 0.016 between anterior STEMI subgroup. No significant difference in inferior wall myocardial infarction subgroups in follow up study, (66.91 ± 4.04) % and (66.44 ± 6.35%) %, p = 0.844. (Table 3)

**Table 3.** Results of LVEF in-hospital and 12-month Follow-up.

| LVEF | Total patients |  |  | Inferior Wall Myocardial Infarction Subgroup |  |  | Anterior Wall Myocardial Infarction Subgroup |  |  |
| --- | --- | --- | --- | --- | --- | --- | --- | --- | --- |
|  | VCR (n=29) | Control (n=31) | P Value | VCR (n=12) | Control (n=11) | P Value | VCR (n=17) | Control (n=20) | P Value |
| Day 3 post PPCI | (57.6 ± 6.2) % | (52.9 ± 8.5) % | 0.018 | (61.25 ± 3.93) % | (61.60 ± 4.57%) % | 0.849 | (55.1 ± 6.3) % | (48.6 ± 6.4%) % | 0.003 |
| 12-month follow up | (64.12 ± 6.35) % | (59.44 ± 8.20) % | 0.025 | (66.91 ± 4.04) % | (66.44 ± 6.35%) % | 0.844 | (62.07 ± 7.06) % | (55.94 ± 6.70) % | 0.016 |

LVEF indicates left ventricular eject fraction; VCR, volume-controlled reperfusion; PPCI, primary percutaneous coronary intervention.

### CMR study

LGE-CMR results on day 5-7 post PPCI indicated no significant difference between VCR group and control group, in proportion of total mass of left ventricular myocardium, regional mass of myocardial infarction, MVO region mass and the MVO region mass/left ventricular myocardial total mass respectively. Neither in patients between the anterior and inferior wall myocardial infarction subgroups. LVEF of VCR and control groups were (48.85 ± 12.61) % vs (46.23 ± 11.61) %, p = 0.483. In anterior wall myocardial infarction subgroup, LVEF were (50.55 ± 7.55) % and (40.53 ± 7.39) %, p = 0.002, between VCR and control group, respectively (Table 4).

**Table 4.** Results of LGE-CMR.

|  | CMR result in total patients |  |  | Inferior Wall Myocardial Infarction Subgroup |  |  | Anterior Wall Myocardial Infarction Subgroup |  |  |
| --- | --- | --- | --- | --- | --- | --- | --- | --- | --- |
|  | VCR (n=21) | Control (n=22) | P Value | VCR (n=7) | Control (n=8) | P Value | VCR (n=14) | Control (n=14) | P Value |
| Total LV mass, g | 120.83<br>(97.70, 135.69) | 117.38<br>(104.74, 136.78) | 0.846 | 113.02<br>(85.45, 140.58) | 117.38<br>(97.69, 127.42) | 0.955 | 122.30<br>(100.96, 135.13) | 116.10<br>(106.65, 139.13) | 0.839 |
| IS mass, g | 29.19<br>(24.01, 48.85) | 38.48 (18.90, 46.30) | 0.981 | 27.19 (18.63, 33.35) | 21.29 (12.78, 29.48) | 0.336 | 42.89 (24.36, 63.66) | 44.01 (35.56, 49.98) | 0.910 |
| MVO zone mass, g | 0.92<br>(0.41, 2.55) | 1.67 (0.00, 3.02) | 0.903 | 0.870 (0.630, 1.730) | 0.000 (0.000, 1.025) | 0.072 | 1.14 (0.18, 3.88) | 2.31 (1.36, 3.51) | 0.511 |
| MVO / LV, mass | 0.010<br>(0.003, 0.236) | 0.012 (0.000, 0.026) | 0.845 | 0.007 (0.006, 0.014) | 0.000 (0.000, 0.009) | 0.072 | 0.011 (0.001, 0.034) | 0.022 (0.008, 0.028) | 0.571 |
| MVO / IS, mass | 0.032<br>(0.010, 0.066) | 0.032 (0.000, 0.079) | 0.714 | 0.051 (0.025, 0.063) | 0.000 (0.000, 0.025) | 0.054 | 0.032 (0.004, 0.109) | 0.062 (0.020, 0.081) | 0.482 |
| CMR-LVEF, % | 48.85 ± 12.61 | 46.23 ± 11.61 | 0.483 | 45.45 ± 19.65 | 56.21 ± 11.15 | 0.207 | 50.55 ± 7.55 | 40.53 ± 7.39 | 0.002 |

VCR: volume-controlled reperfusion; MVO: microvascular obstruction; CMR-LVEF: cardiovascular magnetic resonance-left ventricular ejection fraction; LV: left ventricular; IS: Infarction size

## DISCUSSION

STEMI VCR is a proof-of-concept trial to test the feasibility and safety issue. The main findings of this study were that: VCR was feasible in PPCI practice in patients with acute STEMI; No MACEs reported in VCR group during in-hospital and 12-month follow up; Echocardiography based LVEF reserved better in VCR group, especially in anterior subgroup compared with control group both by in-hospital echo and CMR study; Improved LVEF found in VCR group at 12-month follow-up.

The concept of controlled reperfusion (CR) was originally come from the surgery procedure aim to prevent reperfusion injury by modify perfusate or the mechanical properties, such as pressure and flow. ^7^ The easy way to transfer CR to PPCI procedure in STEMI patient was minimally invasive mechanical intervention, namely, pre-dilation with a small balloon and defer stenting. However, the uncontrolled high-pressure reperfusion and re-occlusion made difficult to maintain sustained low pressure blood flow even in recent GUARD trial.^8^ Gentle reperfusion flow rate achieved by extracorporeal pump during operation or in animal study while few reports in accurate blood flow control in PPCI in clinical scenario. IPost method can be considered as one type of CR with repeated non or all switch by balloon inflating and deflating intra coronary artery.^9^ The main barrier for application of stable gentle reperfusion in PPCI lies in two aspects, to change this non or all switch into intra coronary faucet allowing for accurate and stable flow rates, and the feasibility to set up whole system within minimum time, ideally less than 60 seconds.

Ferrera et al demonstrated low pressure reperfusion can reduce lethal myocardial reperfusion injury even 10-20 mins’ late after the initiation of reperfusion. This study indicated that short time balloon manipulation followed with gentle reperfusion is possible in reperfusion injury prevention.^10^

### Rational and Design of VCR

VCR method might attenuate reperfusion injury in the following aspects: prevent the extravasation of plaque debris, avoid abrupt artery blood restoration, and reduce distal embolization.

Plaque debris and platelet or neutrophil aggregates arise mechanical obstruction of the microcirculation, compromise efficient tissue perfusion. MGuard stent design affording trapping and exclusion of thrombus and friable atheromatous debris before embolization to the distal microvasculature, with superior rates of epicardial coronary flow and complete ST-segment resolution although no significant differences in MVO.^11, 12^ However, TLR rate was higher because of complicated stent design.^12^ We hypothesis that sustained balloon inflation on culprit lesion in VCR, not only to prevent extravasation of plaque debris, also might prevent thrombus embolized distally with antegrade blood flow.

VCR provide stable reperfusion and avoid abrupt or high intra-coronary pressure blood flow through manual injection of diluted artery blood via aspiration catheter at controlled speed and amount. Once reperfusion related decreased heart rate or blood pressure happened, frequently during RCA and LCX recanalization, pause manual injection until all parameter back to stable. There were 20-30 seconds pause waiting for diluted artery blood preparation between each circle that also mimic IPost-like intermittent blood flow. GUARD study indicated pressure-controlled reperfusion of the culprit vessel by means of gradual reopening of occluded infarct-related artery led to better preserved coronary microvascular integrity and smaller myocardial infarction size.^8^

There are uncertainties and risks for thrombus dislocation to side branch or distal part following balloon dilatation. Skyschally et al^13^ believe that microembolization caused by thrombus and plaque debris at reperfusion was existed but the net effect of IPost remains a reduction in infarct size and myocardial infarction expansion on protective effect of reperfusion injury in the presence of microthrombus. Aspiration catheter enrolled in VCR method is mainly served as reperfusion route. Bail out use of aspiration catheter in case of distal thrombus or intra aspiration catheter nitroprusside might be helpful to ensure TIMI III blood flow after VCR.

The establishment of VCR method relied on four key issues: how to set up “intra coronary bypass faucet”, composition of perfusate, the amount and duration of gentle reperfusion.

Coronary seal balloon has clinical application before. In PREPARE study and PROXIMAL study, ^14, 15^ a proximal coronary protection device (The Proxis® device, St. Jude Medical, St. Paul, Minnesota) actively suspended at the proximal part in culprit vessel above culprit lesion, then a balloon catheter and stent advanced through a 3.5F catheter to complete routine PCI manipulation. These trials proved the feasibility and clinical safety for Proxis® device. However, no significant difference found in complete STR at 60 min. Abrupt blood flow restoration might cause reperfusion injury.

Dual-catheter technique is necessary under current coronary intervention equipment condition in Chao-Yang Hospital in this study. VCR procedure need two separate artery route, radial and femoral route, respectively. Coronary pressure monitoring and blood sample preparation via guide catheter with balloon catheter inside, the other guide catheter contended aspiration catheter only. 7F guide catheter can allow one balloon catheter and a micro-catheter inside, but it was difficult to satisfy manual reperfusion volume of 20ml/min via 1.9F micro-catheter over 20mins, neither with over-the-wire balloon. Final or bail out aspiration was necessary when distal thrombus was visualized. Existed femoral route facilitated IABP implantation when needed. Therefore Dual-catheter technique was a reasonable choice in this study.

Perfusate was composed of artery blood taken from guiding catheter and equal amount of heparin normal saline. Heparin normal saline diluted artery blood might help flush out the “metabolic byproduct” in microvasculature in area at risk or infarct core. However, since intramural hemorrhage (IMH) happened based on MVO, we doubted if excessive heparin might exaggerate IMH.^16^ Amier et al show occurrence of IMH was associated with anterior infarction and glycoprotein IIb/IIIa inhibitor treatment.^17^ Minimum amount of heparin in the solution was 20U per ml apart from routine heparin bolus of 70-100U/Kg. Synchronize CMR observation of MVO and IMH will be necessary in stage II RCT study.

Direct volumetric blood flow measurement in selective coronary arteries in conscious humans is a challenging task does not to mention for STEMI patients.^18^ Estimating reperfusion volume is subjective parameter, we estimated minimum volume needed for coronary blood flow at 90% stenosis was 10 ml/min based on 60-80ml/min of blood flow per minute per 100g myocardium in a mid-weight normal person.^18^ Otterspoor et al^19^ have confirmed the safety and feasibility of intracoronary infusion of hypothermia normal saline at a rate of 10-30ml/min. Wang et al^20^ reported a pilot intracoronary hypothermia study in STEMI patients, intracoronary perfusion with a modified DIVER aspiration catheter (MicroPort^®^, China). Perfusion volume initiated from2.5-5ml/min to 10-30ml/min and maintained for 15min. Standard IPost takes about 4-10mins. Hence, VCR reperfusion parameters were set at 20ml (10ml artery blood+10ml heparin normal saline)/min, last for 10 cycles. Total VCR procedure duration was 25-30min including VCR setup and reperfusion. Normally, autoregulation of occluded coronary artery recovered above 30 mins after recanalization.^8^

### Study Analysis

Serial CMR imaging of patients with STEMI in the first few days after PPCI provided evidence that the incidence and extent of MVO vary with time. MVO extent has been reported to peak at 4 to 12 hours, remain stable the first 2 days, and to reduce in size by day 10 post STEMI. Patients who develop adverse LV remodeling had persisting MVO to day 7.^21^ There is no specific exam date for CMR after PPCI. In this study, CMR date was set on day 5 to 7 because some patients had IABP implant to day 3 after PPCI, some patients might in unstable condition. Since this is a small cohort of patients enrolled in this study, and more than 20% of cases in each group failed to complete CMR examination, we could not evaluate left ventricular function by both ultrasound parameters and CMR parameters in each patient.

The occurrence rate of MVO in patients with STEMI ranges from 20% to 60% in acute phase, the highest occurrence was 84%.^22^ In this study, the occurrence of MVO was over 80% in both of VCR and routine PCI group without significant difference. Previous investigation on MVO occurrence seldom describes culprit lesion location, nor the thrombotic load.^16, 23^ Culprit lesion may be scattered in different part in culprit vessel, such as proximal, mid-segment or distal part, resulting in differences in infarct area. The criteria for this study emphasized proximal total occlusion lesions in dominant coronary artery. MVO occurrence rate parallel with infarct area especially in anterior STEMI subgroup.

Backhaus et al^24^ confirms increased mortality for LAD and LCX as compared to RCA lesions and reports IRA-specific underling morphologic as well as function difference following AMI. LV disfunction was higher in LCX and LAD occlusion patients. Proximal LAD occlusion predicts MVO extent in clinical setting. Our results were consistent with this idea. Statistic results show less MVO and IS in RCA related lesion compared with LAD, which consistent with the relationship between MVO and infarct area. We hypothesis a smaller number of patients in LAD subgroup could not show significant difference in MVO as we expected. However, our result suggests that stable condition and better blood flow during PPCI in VCR group might get long-term benefit compared with patients had abrupt antegrade blood restoration in control group.

The reason why there was no difference in MVO between two groups probably related to the following aspects. MVO was a CMR index instead of histological index in this study. LGE-CMR underestimates MVO size by roughly 30%, not mentioned the unknow ratio of hemorrhage, erythrocyte plugging and loss of microvascular integrity. ^25^ Myocardium edema is dynamic pathophysiological process in patient with STEMI and is an important factor attributive to MVO. Myocardial edema during the first week after ischemia reperfusion follows a bimodal pattern.^26^ The first wave happened immediately after reperfusion and dissipates at 24 hours, mainly caused by reperfusion injury. The second wave of edema appears around day 7 after reperfusion, this part of edema probably caused by cell damage and inflammatory procedure.^27^ Consider the time-variant feature of MVO, MVO on day 5-7 post PCI was preferable to reflect myocardial cell damage process but might be affected markley by the second wave of myocardium edema. Myocardial hemorrhage based on MVO is a determinant of MI size which was not observed in this study, myocardium edema and IMH might interfere each other and contribute to MVO.^28^ VCR introduced gentle reperfusion from proximal part of culprit vessel try to have global protection while MVO was normally located in infarct core, means MVO detected by LGE-cMRI might or might not reflect global microvascular damage. Ideally, MVO should be assessed at dynamic time point, combine with physiological assessment, such as IMR and perfusion weighted imaging, to assess microvasculature protection effect and myocardial salvage.

LVEF is a strong predictor of long-term outcome in patients with STEMI in clinical setting. Lower LVEF correlates in-hospital and long-term mortality.^29^ The acute decline in LV function manifests because of transient stunned and hibernation myocardium. However, in-hospital LVEF decline indicate poor prognosis than follow up study in hemodynamic stability. CMR based LVEF is more accurate than echocardiography. Our results show LV function differed in all patients between VCR and control group from in-hospital to 12-month follow up, especially in LAD subgroup. Improved LVEF in VCR group consistent with blood flow and clinical recovery process, favor VCR method might have role reducing reperfusion injury.

The overall operation time course is consistent with routine PPCI. VCR group need less than 15 mins to set up “intra-coronary bypass” perfusion mode and another 15-20 minutes for gentle reperfusion by experienced operator and PPCI team. If the femoral artery route were established, IABP can be prepared simultaneously. Recently, STEMI DTU^30^ was treated with left ventricular unloading 30 minutes before reperfusion, a significant improvement in the ratio of AAR to IS was observed; PiCSO therapy^31^ may improve microvascular function and vasodilatory capacity, which contributes to reducing IS in patients with STEMI undergoing PPCI. PiCSO need 15-30 minutes for sheath preparation and cannulating the coronary sinus before coronary reperfusion. ^32^ Intentionally delaying reperfusion time in STEMI is not recommended but these pilot studies challenge the existing paradigm of STEMI management by reduce myocardial damage and promote myocardial salvage. STEMI VCR shows net benefit of less residual thrombus, reserved TIMI III blood flow and improved heart function inspired the staged II clinical trial.

Current studies of pharmacological targeting of myocardial reperfusion injury, therapies start from peripheral venous or intracoronary after balloon dilation. These procedure and delivery method inevitably lead to prolonged reperfusion time course result in ambiguous results.^3^ Distal quantitative perfusion under prophylactic suspend antegrade blood flow provides a reference for pharmacological conditioning medication delivery in the future.

## Conclusion

STEMI VCR study demonstrated feasibility, clinical safe and improved LVEF with VCR method in patients with STEMI during PPCI. VCR method need stage II randomized clinical trial to evaluate its role in reperfusion injury protection.

### Limitation

This is a pilot trail with limited cases enrolled. More than 20% of cases failed to have CMR examination. Stage II RCT is needed to enroll more cases and careful observation parameter such as IMH or IMR. Dual catheter technique is feasible but complicated in PPCI scenario. A 2.7-2.8F microcatheter might be considered instead of aspiration catheter via single 7F guide catheter, ideally radial artery route only.

## Data Availability

All the data mentioned in the manuscript are available to public.

